# Building case investigation and contact tracing programs in U.S. state and local health departments: a conceptual framework

**DOI:** 10.1101/2023.01.07.23284294

**Authors:** Alexandra Woodward, Caitlin Rivers

## Abstract

The COVID-19 pandemic and earlier health events have demonstrated that when effectively implemented, case investigation and contact tracing (CI/CT) can break chains of transmission by promptly identifying, quarantining, and monitoring the contacts of infected cases, thereby limiting further spread of a disease in a community. Many public health experts agree that implementing CI/CT at the early stages of an outbreak can be an extremely effective approach to controlling an outbreak; as such, health departments must have CI/CT capacities in place prior to the detection of an outbreak to ensure readiness to respond. At the onset of the COVID-19 pandemic, and to this day, U.S. state and local public health departments lack comprehensive CI/CT guidelines that clearly define the capabilities, capacities, outcomes, and impacts of CI/CT programs. This research has resulted in the first comprehensive analysis of the goals, capabilities, and capacities of CI/CT programs, as well as a conceptual framework that represents the relationships between these program components and considerations. Our findings highlight the need for further guidance to assist U.S. state and local public health departments in shifting CI/CT program goals as outbreaks evolve. Moreover, training the public health workforce on making decisions around CI/CT program implementation during evolving outbreaks is critical to ensure readiness to respond to a variety of outbreak scenarios.

## Introduction

Case investigation and contact tracing (CI/CT)* are fundamental public health strategies for controlling and preventing the transmission of infectious diseases. As with any intervention, they are most effective as part of a multifaceted response to an outbreak. When effectively implemented, CI/CT breaks chains of transmission by promptly identifying cases (individuals with probable or confirmed infections) and their contacts (individuals at risk of infection because of their exposure to a known case), notifying contacts of a potential exposure, and making recommendations (such as quarantine) to prevent onward transmission.^1,2^

CI/CT have been used to control transmission in a variety of settings for diseases such as tuberculosis, HIV and during the 2014 Ebola epidemic in West Africa.^3^ Despite CI/CT having been employed to control and mitigate transmission in historic disease outbreaks, the U.S. public health community lacked comprehensive CI/CT guidelines that would enable state and local public health officials to scale-up CI/CT programs at the onset of the COVID-19 pandemic.^4^ Many guidance documents developed during the pandemic focused on explaining how to conduct CI/CT. However, the public health community is still lacking disease-agnostic, comprehensive guidance documents describing what a CI/CT program may be capable of accomplishing during an outbreak and the capacities needed to implement and sustain such a program.

Public health experts have emphasized that implementing CT at the early stages of an outbreak, when incidence is low and during periods of lower transmission, will have the greatest impact on controlling the outbreak. CT may be challenging to maintain in certain epidemiologic situations, such as during periods of widespread, high transmission (e.g., during the Delta phase of the COVID-19 pandemic) because the numbers of cases and contacts that require follow-up exceeds the capacity of even well-staffed CI/CT programs. The ability of a state or jurisdiction to rapidly scale-up CT at the beginning of an outbreak impacts the success of these programs in controlling the outbreak.^3,4^ As such, it is critical to have sufficient CI/CT capacities and capabilities in place at the state and local levels *prior* to the detection of an outbreak to ensure that these programs are ready to be leveraged and deployed when health events are detected.

A CI/CT program consists of **capacities**, which are the organizational, technical, and social resources (such as governance, funding, workforce, and technology) needed to support the program’s **capabilities**. Public health experts have noted that CI/CT programs may offer a variety of capabilities beyond reaching the maximum number of cases and contacts, such as educating the public on the disease and personal risk mitigation measures; generating data that informs our understanding of the disease and connecting people to adjacent public health services.^4-7^ The capabilities are aligned with the goals of conducting CI/CT and refer to what the program can accomplish. The goals are aligned with the **outcomes** of a CI/CT program, which ultimately drive the **impacts** of the program. This research aims to identify these components and build a conceptual framework that describes their interrelatedness (see **Figure 1** for a preliminary conceptual framework). Findings may contribute to guidance documents that assist public health department practitioners and officials in planning robust and impactful CI/CT programs so they are ready to be scaled-up in the early stages of an outbreak.

**Figure 1:**
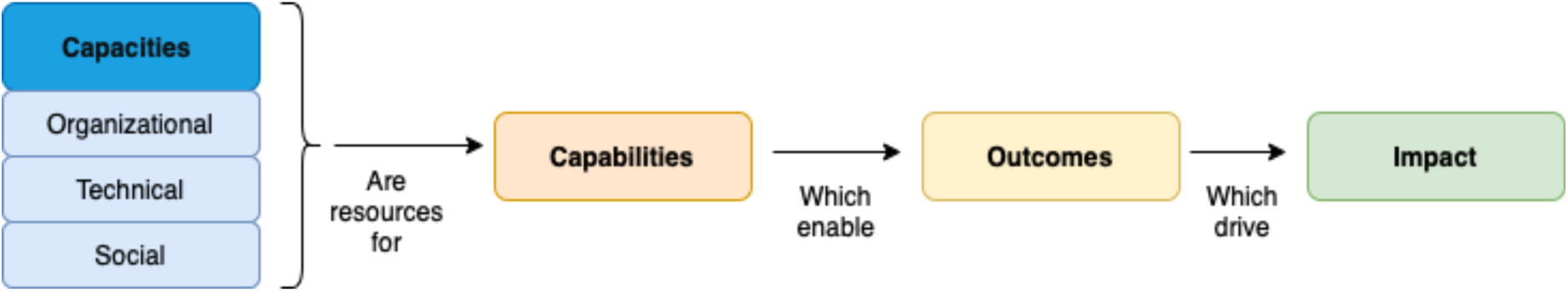
Initial Conceptual Framework for CI/CT Program Capacities, Capabilities, Outcomes, and Impacts.

## Methods

### Research Design Overview

We conducted a narrative literature review to identify the previously documented capacities and capabilities of CI/CT programs. A narrative review is a type of non-systematic literature review aimed at “identifying and summarizing what has previously been published, avoiding duplications, and seeking new study areas not yet addressed.”^8^ A comprehensive, systematic review of scholarly literature was not feasible, as many of the capacities and capabilities of CI/CT programs are described in grey literature such as guidance documents. However, peer-reviewed publications were reviewed using specific criteria to augment our understanding of capacities and capabilities. Once draft lists of CI/CT capabilities and capacities were identified, we conducted qualitative interviews with 10 U.S. state and local public health departments as well as four public health experts to confirm the lists and determine if there were any missing or redundant capacities and capabilities. The interviews also discussed how the capacities support specific CI/CT program capabilities and informed the conceptual framework to describe these relationships.

### Narrative Literature Review

The narrative literature review was conducted in two stages. In Stage 1, we searched Google and Google Scholar for COVID-19 CI/CT guidelines developed by U.S. agencies, international organizations, and nongovernmental organizations for documents published between January 2020 and June 2022. We focused on guidelines developed during the COVID-19 pandemic, as there were limited guidance documents published on CI/CT prior to the pandemic (guidance was primarily focused on how to conduct CI/CT, not a comprehensive understanding of the capacities and capabilities of CI/CT). Moreover, lessons learned around CI/CT from previous outbreaks such as Ebola, sexually transmitted infections, and tuberculosis were incorporated into many of the COVID-19 CI/CT guidelines; many of these lessons were ascertained from peer reviewed literature. Guideline documents met inclusion criteria if they discussed the goals, capabilities, and capacities of a CI/CT program to any extent.

Excluded from the analysis were guidelines produced by non-U.S. countries (as we aimed to focus on CI/CT in U.S. jurisdictions, rather than non-U.S. settings) and guidelines focused solely on providing step-by-step instructions on conducting CI/CT (however, guidelines that incorporated discussion of goals, capabilities, or capacities into the instructions were included). Once all guideline documents were reviewed, we summarized a list of distinct capabilities and capacities. Capacities were bucketed into three broad domains – Organizational, Technical, and Social – based on the nature of their function.

In Stage 2, we searched peer-reviewed publications and grey literature published between January 1970 and June 2022 using SCOPUS and Google Scholar to supplement our understanding of CI/CT capabilities and capacities. Inclusion criteria included documents or articles that addressed: CI/CT models and approaches; the implementation of CI/CT programs; secondary outcomes or unintended consequences of CI/CT; lessons learned and recommendations for CI/CT programs; discussions around the capacities identified from Stage 1 in the organizational, technical, and social domains, and publications from U.S. state and territories only. Excluded from the analysis were publications focused solely on quantitative metrics of CI/CT programs (e.g., studies focused on reporting on CI/CT programmatic and performance metrics, such as proportion of community contacts traced) and those focused solely on the development and use of mobile applications for CI/CT. We used key terms from the capacities identified in the guideline documents (Stage 1), as well as broader terms such as “case investigation,” “contact tracing,” and “partner notification.”

### Qualitative Public Health Department and Expert Interviews

Qualitative interviews with U.S. state and local public health departments as well as public health experts were conducted to confirm and elaborate on the goals, capabilities and capacities identified in the narrative literature review. As CI/CT program implementation varied considerably across the U.S. during the COVID-19 pandemic, conducting case studies provides real-world insight into CI/CT programs. A holistic multiple case design was employed, allowing for the conduct of a within- and cross-case comparison of CI/CT programs, each constituting a single unit of analysis.^9^ The strength of this design is that it allows for the development of an in-depth description of each CI/CT program and to identify patterns of similarity or variation between programs.

Participants were selected to represent a diversity of perspectives across two dimensions: sufficient workforce capacity and CI/CT program model type as of December 2020. The first criterion (workforce capacity) was chosen based on a report published by the National Association of County and City Health Officials (NACCHO) in April 2020 that called for a massive expansion of professionals and trained volunteers to conduct CI/CT across the country. The report recommended that 30 professionals per 100,000 population, distributed across health departments in an equitable fashion (using a per capita formula so health departments that serve smaller populations have sufficient workforce capacity), were needed to effectively conduct CI/CT during emergency situations such as the COVID-19 pandemic.^10^ This workforce recommendation assumed, at the time, that a sufficient number of investigators and tracers per population would prevent widespread community transmission. It was also one of the only quantitative measures of CI/CT program capacities in every U.S. state in 2020 and 2021. Using the 30 per 100,000 population benchmark, state participants were selected from data collected in a December 2020 survey by the Johns Hopkins Center for Health Security and National Public Radio to include both participants that did and did not meet the benchmark.^11^ This selection criteria assumes that the workforce benchmark of 30 per 100,000 population was indeed sufficient for effective CI/CT in a state or jurisdiction.

The second dimension for participant selection was CI/CT model type, of which there are three: first, the “In-House” model is where state and local officials lead the CI/CT programs, hiring or recruiting volunteers as needed; second, the “Contracting” model is where the state contracts with a company or organization for CI/CT work/hiring, and third, the “Partnering” model involves the state leading the efforts but relies on partners for training/staffing. To explore a variety of participants employing different models, all models were represented in CI/CT program participants selected, either individually or as a combination of models, as many states combined models and approaches may have changed throughout the pandemic.^12^ To achieve greater diversity and representation across the country, several additional factors in participant selection were considered: the Governor’s political party as of December 2020, the U.S. census region, and population density (based on 2022 U.S. census data) in the public health department’s jurisdiction. We aimed to have representation for both political parties and all U.S. census regions across the sample; we also aimed to have representation from jurisdictions with both high and low population densities.

The Johns Hopkins Bloomberg School of Public Health Institutional Review Board (IRB) reviewed and designated study protocol (IRB No. 00019635) “exempt” and “not human subjects research.” We recruited participants via outreach supported by a non-profit, professional association that works to advance public health and workforce capacity in state and local jurisdictions in the U.S. Contact information was provided by the organization and a recruitment email with background regarding the study was sent out to a total of seven state public health departments selected using the participant selection criteria, and five agreed to participate. Among the five, three facilitated recruitment to one or two local health departments within the state to participate in the interviews as well. Ten state and/or local health departments in five states participated in the interviews (**Table 1**). A total of 13 interviews were conducted with 15 interviewees (two health departments included more than one interviewee). Interviewees included CI/CT program leadership and senior-level public health staff who managed some or all aspects of a CI/CT program. A snowball approach was used to identify four public health experts in CI/CT to participate in individual interviews from the Association of State and Territorial Health Officials (ASTHO), the Council of State and Territorial Epidemiologists (CSTE), NACCHO, and an advisor to a Health and Human Services (HHS) agency. We deemed that the interviews reached saturation once they failed to uncover any new findings related to the goals, capabilities, and capacities.

**Table 1:**
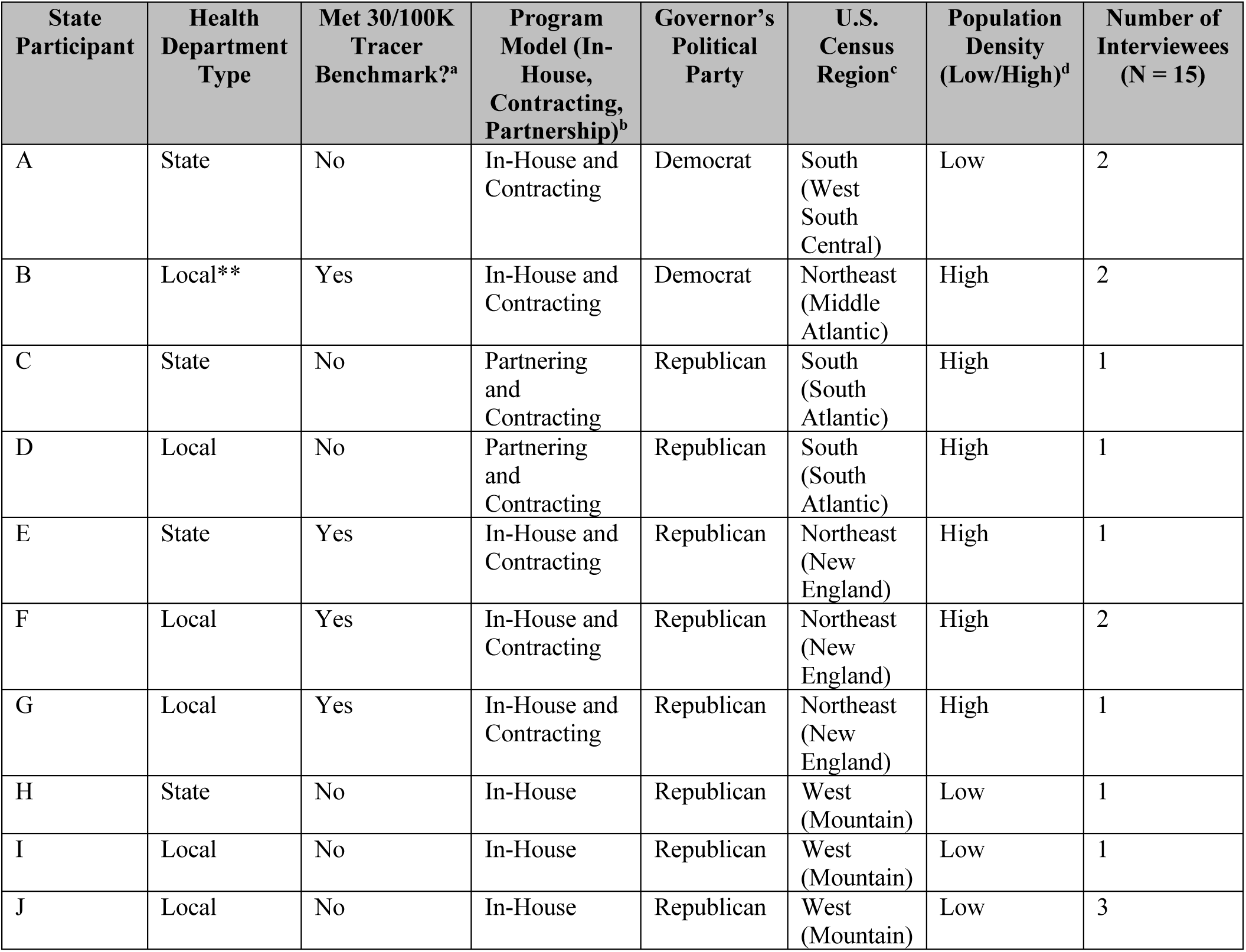
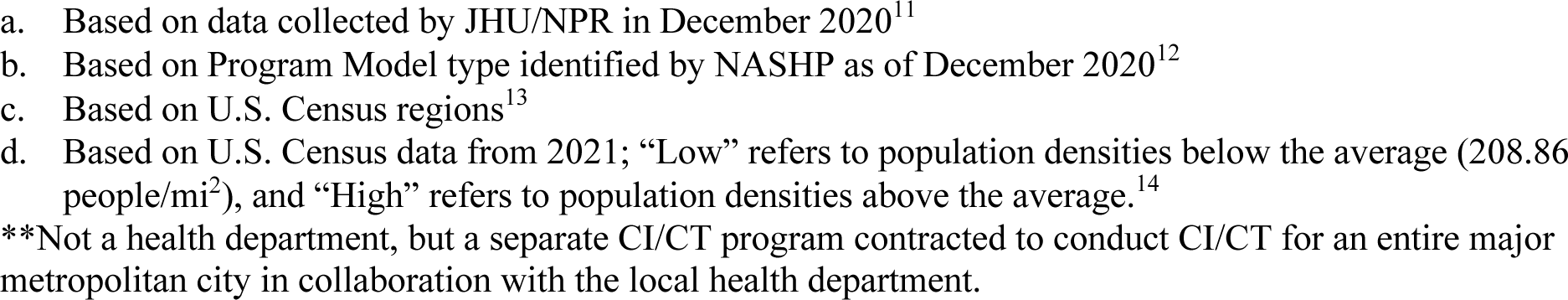
Characteristics of the 10 Participating Health Departments.

| State Participant | Health Department Type | Met 30/100K Tracer Benchmark?<sup>a</sup> | Program Model (In-House, Contracting, Partnership)<sup>b</sup> | Governor's Political Party | U.S. Census Region<sup>c</sup> | Population Density (Low/High)<sup>d</sup> | Number of Interviewees (N = 15) |
| --- | --- | --- | --- | --- | --- | --- | --- |
| A | State | No | In-House and Contracting | Democrat | South (West South Central) | Low | 2 |
| B | Local** | Yes | In-House and Contracting | Democrat | Northeast (Middle Atlantic) | High | 2 |
| C | State | No | Partnering and Contracting | Republican | South (South Atlantic) | High | 1 |
| D | Local | No | Partnering and Contracting | Republican | South (South Atlantic) | High | 1 |
| E | State | Yes | In-House and Contracting | Republican | Northeast (New England) | High | 1 |
| F | Local | Yes | In-House and Contracting | Republican | Northeast (New England) | High | 2 |
| G | Local | Yes | In-House and Contracting | Republican | Northeast (New England) | High | 1 |
| H | State | No | In-House | Republican | West (Mountain) | Low | 1 |
| I | Local | No | In-House | Republican | West (Mountain) | Low | 1 |
| J | Local | No | In-House | Republican | West (Mountain) | Low | 3 |

- a. Based on data collected by JHU/NPR in December 2020<sup>11</sup> - b. Based on Program Model type identified by NASHP as of December 2020<sup>12</sup> - c. Based on U.S. Census regions<sup>13</sup> - d. Based on U.S. Census data from 2021; “Low” refers to population densities below the average (208.86 people/mi<sup>2</sup>), and “High” refers to population densities above the average.<sup>14</sup> - \*\*Not a health department, but a separate CI/CT program contracted to conduct CI/CT for an entire major metropolitan city in collaboration with the local health department.

We developed a semi-structured interview questionnaire to guide the interviews, which lasted one-to-two hours for each participant. All interviews were conducted by one researcher between May and August 2022. Participants received read-ahead materials, which included an overview of the study and the lists and descriptions of capacities and capabilities. The interviewees were given at least one week to review the lists in the read-ahead material prior to each interview and were asked to come prepared to comment on the capacities and capabilities, identifying any missing components or share feedback on their program’s approaches to scaling up capacities to accomplish the capabilities. The interviews were recorded, and the audio recordings were transcribed and coded with inductive approaches based on the draft capacity and capability lists using NVivo 11 software.

## Results

### Narrative Review of Literature

During Stage 1, 17 Guideline documents (**Table 2**) published between January 2020 and June 2022, met the inclusion criteria, were reviewed, and goals, capabilities and capacities were documented in draft tables. All Guidelines included discussion of both capabilities and capacities to varying degrees (**Tables 3 and 4**). In Stage 2 of the narrative review of literature, 94 peer-reviewed publications met the inclusion criteria and were included in the analysis to provide additional insight into the goals, capabilities, and capacities identified in the Guideline documents.

**Table 2:**
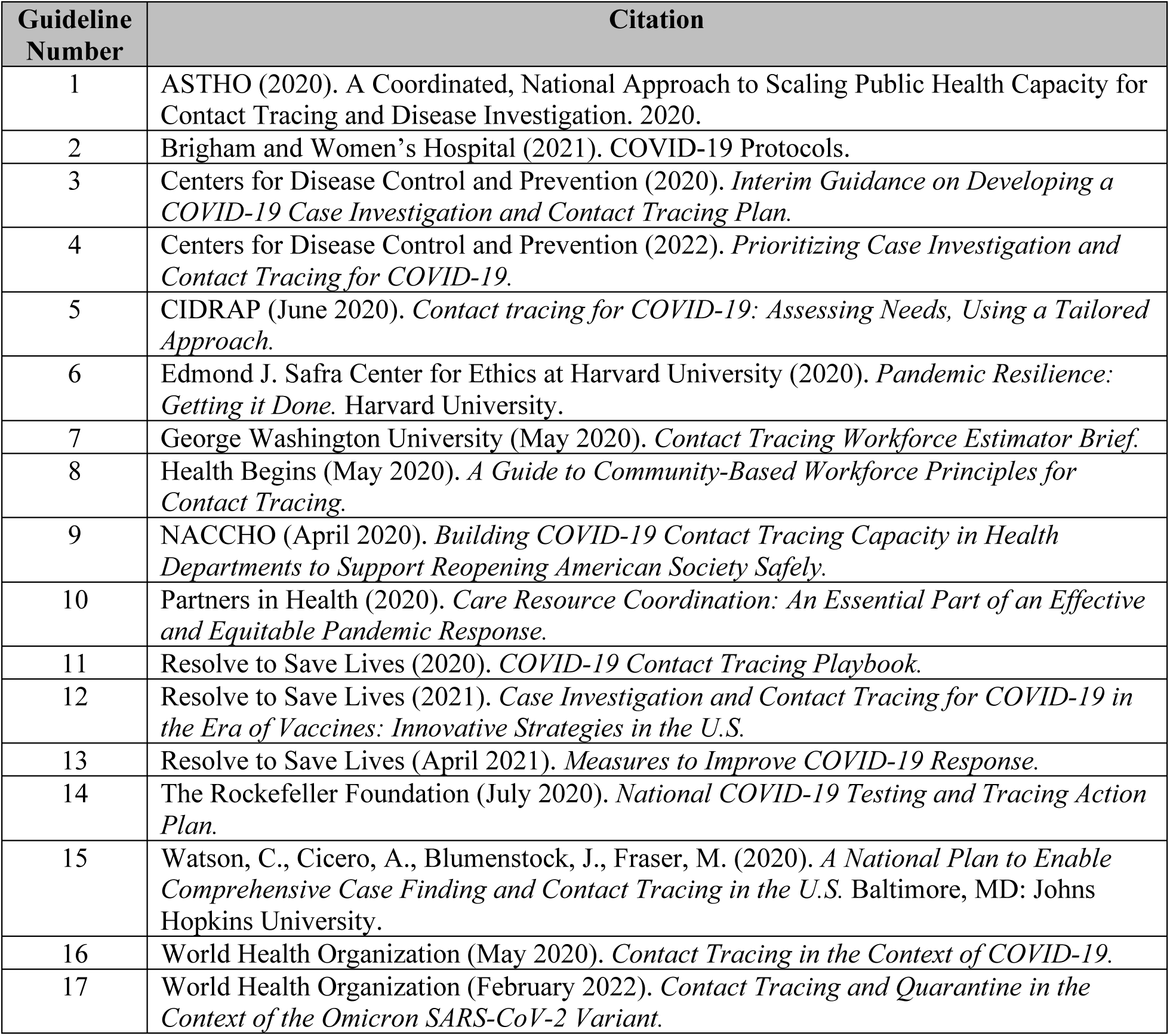
COVID-19 CI/CT Guideline and Recommendation Documents (N = 17)

| **Guideline Number** | **Citation** |
| --- | --- |
| 1 | ASTHO (2020). A Coordinated, National Approach to Scaling Public Health Capacity for Contact Tracing and Disease Investigation. 2020. |
| 2 | Brigham and Women’s Hospital (2021). COVID-19 Protocols. |
| 3 | Centers for Disease Control and Prevention (2020). *Interim Guidance on Developing a COVID-19 Case Investigation and Contact Tracing Plan*. |
| 4 | Centers for Disease Control and Prevention (2022). *Prioritizing Case Investigation and Contact Tracing for COVID-19*. |
| 5 | CIDRAP (June 2020). *Contact tracing for COVID-19: Assessing Needs, Using a Tailored Approach*. |
| 6 | Edmond J. Safra Center for Ethics at Harvard University (2020). *Pandemic Resilience: Getting it Done*. Harvard University. |
| 7 | George Washington University (May 2020). *Contact Tracing Workforce Estimator Brief*. |
| 8 | Health Begins (May 2020). *A Guide to Community-Based Workforce Principles for Contact Tracing*. |
| 9 | NACCHO (April 2020). *Building COVID-19 Contact Tracing Capacity in Health Departments to Support Reopening American Society Safely*. |
| 10 | Partners in Health (2020). *Care Resource Coordination: An Essential Part of an Effective and Equitable Pandemic Response*. |
| 11 | Resolve to Save Lives (2020). *COVID-19 Contact Tracing Playbook*. |
| 12 | Resolve to Save Lives (2021). *Case Investigation and Contact Tracing for COVID-19 in the Era of Vaccines: Innovative Strategies in the U.S.* |
| 13 | Resolve to Save Lives (April 2021). *Measures to Improve COVID-19 Response*. |
| 14 | The Rockefeller Foundation (July 2020). *National COVID-19 Testing and Tracing Action Plan*. |
| 15 | Watson, C., Cicero, A., Blumenstock, J., Fraser, M. (2020). *A National Plan to Enable Comprehensive Case Finding and Contact Tracing in the U.S.* Baltimore, MD: Johns Hopkins University. |
| 16 | World Health Organization (May 2020). *Contact Tracing in the Context of COVID-19*. |
| 17 | World Health Organization (February 2022). *Contact Tracing and Quarantine in the Context of the Omicron SARS-CoV-2 Variant*. |

**Table 3:** CI/CT Goals and Capabilities Referenced in Guideline Documents.

| **CI/CT Goals** | **CI/CT Capabilities to Support Goal** | **Number of References**<br>**(N = 17**<br>**Guidelines) (%)** |
| --- | --- | --- |
| **Goal 1: Isolate and quarantine cases and contacts** | Identify cases and contacts | 11 (65%) |
|  | Alert cases that they should isolate and elicit contacts | 11 (65%) |
|  | Alert contacts that they should monitor for symptoms, get tested, quarantine, alert their own contacts | 11 (65%) |
|  | Monitor cases and contacts for symptoms and isolation/quarantine adherence | 11 (65%) |
|  | Conduct source investigation (backward CT) | 5 (29%) |
|  | Connect cases/contacts to care, prophylaxis, treatment, out-of-home isolation/quarantine, and wraparound services | 11 (65%) |
| **Goal 2: Educate cases and contacts** | Educate cases/contacts on the disease (clinical, epidemiologic, risk factors, etc.), using best practices to counter misinformation | 7 (41%) |
|  | Educate cases/contacts on the importance and availability of vaccination, prophylactics, and therapeutics, using best practices to counter misinformation | 6 (35%) |
|  | Educate cases/contacts about on the importance of CI/CT and strategies for notifying their own contacts | 5 (29%) |
|  | Promote the use of digital tracing tools such as apps and SMS | 3 (18%) |
| **Goal 3: Build community trust** | Build trust in communities by countering misinformation about the utility and processes of CI/CT to improve participation in CI/CT and compliance with isolation, quarantine, and other efforts to mitigate the disease transmission | 6 (35%) |
|  | Enable resiliency of communities by connecting cases and contacts to a range of referrals for longer-term support, such as crisis and recovery counseling | 3 (18%) |
| **Goal 4: Use CI/CT data to inform decision-making** | Inform shifts in CI/CT priorities and strategies, such as prioritizing high-risk and vulnerable populations if program capacities are stretched or limited | 3 (18%) |
|  | Inform the natural history and epidemiology of the disease | 2 (12%) |
|  | Inform public health decision-making around containment and mitigation measures | 3 (18%) |
| **Goal 5: Provide adjacent public health services** | Serve as an entry point into clinical and public health systems | 2 (12%) |

**Table 4:** CI/CT Program Capacities Referenced in Guideline Documents.

| Domain | Capacity | Number of References<br>(N = 17 Guidelines) (%) |
| --- | --- | --- |
| Organizational | Governance | 5 (29%) |
|  | Funding*** | 8 (47%) |
|  | Partnerships | 9 (53%) |
|  | Workforce | 15 (88%) |
| Technical | CI/CT Processes, Protocols and Forms | 11 (65%) |
|  | CI/CT Data Collection, Management, and Analysis Systems | 10 (59%) |
|  | Digital and Technology Solutions | 10 (59%) |
|  | Privacy and Data Sharing | 8 (47%) |
|  | Metrics and Monitoring | 6 (35%) |
|  | Clinical Consultation | 6 (35%) |
|  | Procurement and Distribution of Materials | 3 (18%) |
|  | Out-of-Home Isolation/Quarantine | 4 (24%) |
| Social | Wraparound Support Services | 12 (71%) |
|  | Public Communication and Engagement | 10 (59%) |

\*\*\* Added after case study interviews were complete, then re-read the Guideline documents to document discussions around Funding.

#### CI/CT Goals and Capabilities

**Table 3** includes the Guideline documents’ frequency of referencing each capability; capabilities were aligned with each goal, as each capability supports specific goals (however, as discussed later in the conceptual framework, capabilities may support multiple goals or outcomes). The majority of the Guideline documents were focused on Goal 1 (to isolate and quarantine cases and contacts). This finding is not surprising, as the primary, documented goal of CI/CT is to isolate and quarantine cases and contacts. Goals 2 – 5 and their supporting capabilities were documented less frequently in the Guideline documents.

#### CI/CT Capacities

The capacities ascertained from the review of literature are described in **Table 4**, which includes the Guideline documents’ frequency of referencing each capacity. The majority of the Guideline documents referenced the importance of several CI/CT capacities: Partnerships; Workforce; CI/CT Processes, Protocols, and Forms; CI/CT Data Collection, Management and Analysis Systems; Digital and Technology Solutions; Wraparound Support Services, and Public Communication and Engagement. Less than half of the Guideline documents discussed Funding; Privacy and Data Sharing; Metrics and Monitoring; Clinical Consultation; Procurement and Distribution of Materials, and Out-of-Home Isolation/Quarantine. As several of these capacities were discussed during the Qualitative Interviews (discussed below), future Guideline documents should incorporate these elements and address their implementation and scale-up.

### Qualitative Public Health Department and Expert Interviews

Once the lists of goals, capabilities, and capacities were finalized based on the narrative literature review, they were presented to interviewees, who confirmed the lists and provided greater detail and nuance to the goals, capabilities and capacities. The lists presented to the interviewees did not include the number of references from each Guideline document to reduce any biases in the discussion of goals, capabilities, and capacities. Tables 2 and 3 underwent slight revisions based on responses from the case study interviews, and interview responses informed the descriptions of each capacity (included below).

#### Capabilities

Interviewees confirmed that all goals and capabilities (**Table 3**) are critical with respect to containing and/or mitigating outbreaks, but goals may vary depending on the disease scenario. For example, a health department may focus on Goals 1 and 2 during an outbreak of a pathogen with high infectivity, pathogenicity, and lethality (such as smallpox), especially if capacities are limited. Alternatively, if not conducting universal CI/CT during a COVID-19 outbreak, a health department try to accomplish Goal 1 for vulnerable and at-risk populations, while dedicating capacities towards Goals 2-5 as well. In addition to confirming the goals and capabilities in **Table 3**, several interviewees discussed cross-cutting themes regarding the goals and capabilities:

##### Shifting CI/CT goals during outbreaks

Several interviewees noted that as an outbreak evolves and the transmission dynamics of a disease are better understood over time, the goals of a CI/CT program may shift. For example, CI/CT goals around isolating and quarantining cases and contacts changed based on an evolving reproduction number during the COVID-19 pandemic (e.g., many jurisdictions moved away from universal CI/CT during the Delta phase of the pandemic and pivoted to providing education to cases on how to alert their own contacts). Interviewees, particularly those representing local health departments, mentioned that this shift resulted in changing procedures and the need to frequently update training materials and re-train the workforce. When CI/CT goals shift, it is challenging for health departments to rapidly pivot to prioritizing different capabilities to meet the shifting goals. A few sets of guidelines were developed during the COVID-19 pandemic to aid states and local jurisdictions in pivoting their CI/CT goals and practices during varying transmission scenarios ^15,16^

##### Educating Cases and Contacts and Building Community Trust

Given unlimited resources, the participants discussed the importance of demonstrating all CI/CT program capabilities in any disease scenario. However, several participants emphasized that the education of cases and contacts about the disease, the importance and availability of vaccination, prophylactics, and therapeutics, and the importance of participation in CI/CT are critical capabilities that should be prioritized in any scenario. Several interviewees noted that information provided should be actionable at the individual level; should be able to offer an outcome to cases and contacts (e.g., how to access prophylactics, treatment, and wraparound services) and should not be punitive. Building a rapport with cases and contacts to serve as a trusted resource for information and resources for recovery and resiliency improves public trust, which contributes to reaching all other CI/CT goals.

#### Capacities

Many participants noted that while the capacity list was all-encompassing and all capacities are critical to support a CI/CT program, it is not feasible to maintain all capacities at the same time given existing health department funding levels and resources. Interviewees shared their insights on specific capacities and added further detail to assist in the development of definitions below. For example, all interviewees commented on Workforce as being a critical capacity for CI/CT programs, but many added greater detail, such as the need to provide mental health support to the CI/CT workforce during periods when their capacity is stretched thin. The descriptions of the capacities (below) were developed based on both the narrative review of literature and the interviews.

## Organizational Capacities

- **Governance:** A functional public health structure is in place that enables effective and consistent coordination and communication between state and local health departments as well as between health departments and legislators. There is support from state and local legislators, senior health officials and public health leadership for the provision of CI/CT capacities and associated policies.
- **Funding:** There is sufficient permanent, flexible funding that is readily accessible to both state and local health departments to develop, implement, and sustain capacities for CI/CT. Funding amounts by jurisdiction may be determined by models based on population data and community needs. During outbreaks, funding dedicated to CI/CT programs should be based on cost-benefit analyses that consider all CI/CT program capacities, the epidemiology of a disease, and vulnerable/at-risk populations within communities.
- **Partnerships:** Collaboration with a variety of stakeholders is critical to the impact of CI/CT on communities: state and local health departments (within and across states), other relevant government agencies, healthcare providers, public health and clinical laboratories, social services, schools/daycares, congregate settings, community-based organizations, religious organizations, licensing organizations, and federal health agencies.
- **Workforce:** There is rapid and efficient recruitment, training, deployment, and retention of case investigators, contact tracers, information technology (IT) staff, and resource support staff, managed by a sufficient program and supervisory staff that can ensure the quality of the workforce. Federal, cross-state, and/or in-state reach-back support can be leveraged to assist with CI/CT as needed. The workforce should be able to create rapport with appropriate language skills, cultural competence, address concerns and barriers to contact elicitation or isolation/quarantine and make connections to appropriate housing and wraparound services. CI/CT workforce should have access to benefits that ensure work-life balance, such as sufficient pay, benefits, and mental health resources.

## Technical Capacities

- **CI/CT Processes, Protocols, and Forms:** CI/CT processes are integrated with existing or new disease surveillance system(s). There are clear processes, protocols and forms for conducting CI/CT in various epidemiologic scenarios (outbreaks, congregate settings, travel), and processes for isolation and quarantine. Staff are able to rapidly update processes, protocols, and forms based on changing federal and state health agency guidance and changing CI/CT practices. Processes are in place to connect cases and contacts with testing, prophylaxis, treatment, and wraparound services.
- **CI/CT Data Collection, Management, and Analysis Systems:** The CI/CT program uses nimble, flexible CI/CT data systems that are interoperable with laboratory, clinical, and other relevant data reporting systems to facilitate real-time data management and reporting. Data systems should integrate telephone software (or other relevant data collection technology) and contain data collection scripts and packages that can be rapidly updated by CI/CT staff. The data system should have sufficient support from an IT workforce capable of providing rapid system updates and should ensure best practices for data privacy and security.
- **Digital and Technology Solutions:** Digital products and solutions, such as apps and SMS communications with cases/contacts, are available as needed to supplement manual CI/CT programs. Strategies to employ digital solutions should be in place, including assistance with decision-making around when and where to employ the solutions and how to build public trust around the solutions. Partnerships with the private sector may be needed for development and implementation.
- **Privacy and Data Sharing:** Data reporting processes must abide by privacy and security laws, norms, and standards. Training and certification on data privacy for all case investigators and contact tracers should readily accessible. Data Sharing Agreements are in place and/or can be rapidly facilitated to ensure appropriate access to data within and across state and local health departments, as well as with appropriate federal agencies, to inform decision-making.
- **Metrics and Monitoring:** A plan is in place to monitor and assess CI/CT activities and outcomes. Health departments have the ability to share these metrics with health department or local/state/federal government leadership to inform decision-making.
- **Clinical Consultation:** A plan is in place to connect cases with healthcare providers for clinical consultation in inpatient, outpatient, and telehealth settings. Ensure CI/CT staff are appropriately trained on recognizing clinical manifestations of severe disease that warrant a referral to a clinician. Ensure healthcare providers are familiar with CI/CT and are empowered to encourage patients to respond to CI/CT outreach. Maintain relationships with infectious disease specialists who can inform CI/CT strategies and approaches.
- **Procurement and Distribution of Materials:** There is a sufficient supply of CI/CT-related materials, such as personal protective equipment (PPE), hardware and software, as well as consistent, reliable internet access in both office and remote work settings.
- **Out-of-Home Isolation/Quarantine:** Alternative housing and transportation to alternative housing is available for those who cannot isolate/quarantine at home. Sufficient and trained workforce is in place to connect cases/contacts to alternative housing.

## Social Capacities

- **Wraparound Support Services:** Resources are available to improve ability to comply with isolation and quarantine, such as food, paid sick leave, medication delivery, health insurance, rent/utility assistance, mental health support, and other support services. Decisions around availability of specific resources should be informed by understanding of community-specific needs. Local engagement with relevant organizations, cross-agency and cross-sector partnerships, and sufficient workforce are critical to ensure appropriate resource provision.
- **Public Communication and Engagement:** Public communication and engagement through effective, targeted public messaging to ensure participation in CI/CT and adherence to recommendations is critical. Appropriate communication strategies are implemented for various language-speaking, hard-to-reach, and vulnerable populations. Strategies should be adaptable to changing guidance and should consider ways to offer information, preventive measures, services, and care to cases and contacts.

### Additions and Removals of Capacities based on Interview Feedback

A few capacities were added or removed based on feedback from the qualitative interviews. First, “Testing and Laboratory Services” was a capacity that was initially included from the narrative literature review but was removed based on feedback from the interviews. While the ability to rapidly test cases and contacts and deliver results to public health agencies is a critical operation in conducting CI/CT, testing is part of a broader enabling environment that facilitates more effective CI/CT. Moreover, testing is used for purposes beyond CI/CT, such as therapeutically as a diagnostic tool, for surveillance activities, and to determine population immunity (antibody testing).^17^ As such, testing was removed from the Capacity list, but is included in the broader Enabling Environment as depicted in the conceptual framework (**Figure 2**). Moreover, while “Funding” was not initially identified as a capacity in the narrative literature review, the availability of sufficient funding was discussed extensively during the interviews and was therefore added as a standalone Organizational capacity. The Guideline documents were reviewed once again to collect the frequency of the discussion of funding.

**Figure 2:**
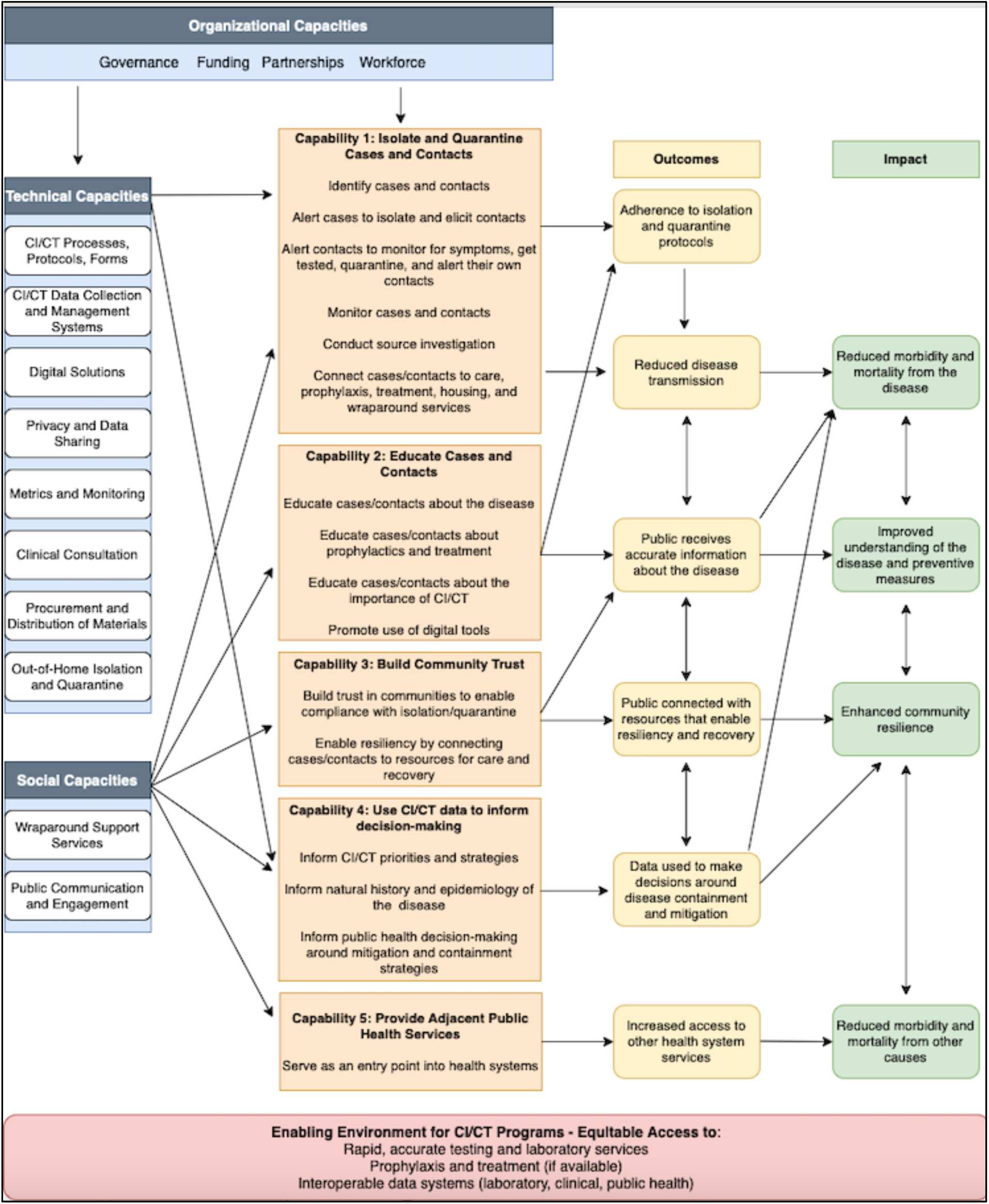
CI/CT Conceptual Framework.

### Conceptual Framework

While the capacities and capabilities are identified in disparate CI/CT guidelines and academic and grey literature, there is no documentation of how the capacities and capabilities relate to each other and the outcomes or impacts of CI/CT. A conceptual framework (**Figure 2**) was developed to visualize the relationship between the capacities, capabilities, outcomes, and impacts of CI/CT. The capacities serve as resources for the capabilities, which support the outcomes (synonymous with the goals in **Table 3**), which drive the impact of CI/CT programs. Based on the narrative literature review and the qualitative interviews, CI/CT programs may ultimately result in the following impacts: (1) reduced morbidity and mortality from a disease; (2) an improved understanding among community members of the disease and preventive measures; (3) enhanced community resilience to outbreaks, and (4) reduced morbidity and mortality from other causes. While not all of these impacts may be realized in all outbreak scenarios, they should all be considered when establishing the goals of a CI/CT program.

Both the narrative literature review and the qualitative interviews indicated that capacities and capabilities are not mutually exclusive; multiple capacities may support more than one capability, which may result in more than one outcome and impact. For example, the Organization and Social capacities (seen in blue) must be in place to support all five capabilities (seen in orange), whereas the Technical capacities support two capabilities. This finding is notable, as a great deal of attention during the COVID-19 pandemic was paid to CI/CT digital solutions such as proximity tracing applications. For example, hundreds of studies on digital solutions were published in peer-reviewed journals during the pandemic, with much less attention paid to many of the Organizational and Social capacities as well as other Technical capacities in terms of the quantity of publications.^18^

Another notable finding from this Framework is the bidirectional relationship between several of the outcomes and impacts. For example, reduced transmission; the public receiving accurate information about the disease; their connectedness to resources that enable resiliency and recovery, and data used to make decisions around containment and mitigation are all inter-related outcomes that rely on capabilities 1-4 to come to fruition. This finding speaks to the need for health departments to strategize the use of multiple capacities to support several capabilities at once, and these capabilities may vary depending on the epidemiology and biology of the pathogen causing the outbreak.

Finally, there are several “Enabling Environment” factors that support the capacities and capabilities: rapid, accurate testing and laboratory services; prophylaxis and treatment (if available); and interoperable data systems to enable the exchange of laboratory, clinical, and public health data.

## Discussion

The identification of the goals, capabilities, and capacities of CI/CT programs, as well as a conceptual framework that describes the relationship between these elements with the outcomes and impacts of CI/CT programs, may guide public health leaders and officials in maintaining and sustaining these programs in “pandemic peacetime” as well as scaling them up during health emergencies. However, the identification of these factors alone is not sufficient to drive the changes needed to better prepare state and local health departments for conducting CI/CT in future outbreaks. The recommendations below describe how this research can be operationalized in the near-term.

### Developing disease-agnostic guidance to address shifting CI/CT goals, capabilities, and capacities during outbreaks

The goals of CI/CT programs may shift during an outbreak. Health departments need to be nimble enough to pivot program capabilities to respond to rapidly changing goals. The U.S. Centers for Disease Control and Prevention and/or public health partners such as ASTHO, NACCHO, and CSTE should develop disease-agnostic guidance on adjusting CI/CT goals and capabilities during an evolving outbreak (as well as when multiple outbreaks are occurring simultaneously); this guidance should be capable of being tailored to disease-specific scenarios.

### Incorporating cost-benefit analysis into guidance documents

The disease-agnostic guidance should incorporate methods to conduct cost-benefit analysis to determine which CI/CT goals may be most cost-effective and impactful given the information available about the epidemiology of the disease. For example, a cost-benefit analysis may conclude that Goals 1 and 5 would optimize a health department’s resources to sufficiently suppress community transmission. Several studies have explored methods and models for determining the cost-effectiveness threshold of investing in and conducting CI/CT during outbreaks; however, further study in this area is needed.^19-21^ Additionally, these methods and models need to be available to public health department leaders so they can be rapidly leveraged at the onset of an outbreak.

### Incorporating decision-making into preparedness exercises

Public health department leaders and staff should demonstrate the ability to pivot capabilities and capacities based on shifting CI/CT goals in different outbreak scenarios. Beyond developing further guidelines, incorporating decision-making around CI/CT goals, capabilities, and capacities into public health preparedness training and exercises is critical to ensuring that these skills are tested and maintained. Several interviewees commented on the need to develop scenario-based practices to strategize CI/CT capabilities and capacities, such as during “high” threat scenarios such as a smallpox outbreak, more “moderate” threat scenarios such as COVID-19, and “low” threat scenarios such as sexually transmitted infections. These scenarios may be tested through routine outbreak response exercises conducted by state and local health departments, such as those required for public health department accreditation by the Public Health Accreditation Board.^22^ Additionally, training for health department staff should include concepts of CI/CT goals, capabilities, and capacities, as well as different health threat scenarios to enable trainees to test their decision-making skills.

Our analysis included several limitations. First, the narrative review of literature excluded peer-reviewed and grey literature publications from outside of the U.S. and literature discussing CI/CT prior to 1970. Future research into the goals, capabilities, and capacities of CI/CT programs should explore programs conducted in non-U.S. countries and multinational efforts, both prior to and after 1970. CI/CT is not a strategy unique to the U.S.; many countries employed CI/CT during the COVID-19 pandemic as well as during previous outbreaks such as Ebola, and lessons learned around these non-U.S. programs may further inform the goals, capabilities, capacities and the conceptual framework. Moreover, while only 10 health departments and four experts from public health organizations were interviewed as part of this research, the sample selection included a broad representation of health departments across the country and public health officials and personnel from both the state and local levels. Few in-depth interviews of health department staff, specific to CI/CT during the COVID-19 pandemic, have occurred to date, and further discussion with additional jurisdictions may yield additional lessons learned.^23^ Furthermore, a subsequent publication will discuss lessons learned from the qualitative interviews around scaling-up and maintaining CI/CT programs.

## Conclusion

This research represents the first comprehensive analysis of the goals, capabilities, and capacities of CI/CT programs in U.S. state and local settings in response to the COVID-19 pandemic. This review enabled the development of a conceptual framework that represents the relationships between capacities, capabilities, outcomes, and impacts of CI/CT programs. The results of this research identified the need for further guidance for state and local public health agencies regarding approaches to shifting CI/CT program goals as outbreaks are detected and evolve, as well as training and testing the public health workforce on decision-making around CI/CT program implementation during varying outbreak scenarios.

* While these processes can be conducted separately with distinct goals and benefits (e.g., case investigation is sometimes conducted without contact tracing) this research refers to these processes as one concept under the term “CI/CT.”^15^

## Data Availability

All data produced in the present study are available upon reasonable request to the authors

## Acknowledgements

We would like to thank our state and local public health department and expert interview participants for sharing their perspectives for this research, as well as 1 anonymous reviewer for their important contributions to this article.

